# Statins Are Associated with Improved 28-day Mortality in Patients Hospitalized with SARS-CoV-2 Infection

**DOI:** 10.1101/2021.03.27.21254373

**Authors:** Zoe N. Memel, Jenny J. Lee, Andrea S. Foulkes, Raymond T. Chung, Tanayott Thaweethai, Patricia P. Bloom

**Affiliations:** Harvard Medical School, Boston, MA; Department of Medicine, Massachusetts General Hospital, Boston MA; Department of Biostatistics, Harvard T.H. Chan School of Public Health, Boston, MA; Liver Center, Division of Gastroenterology, Massachusetts General Hospital, Boston, MA; Biostatistics Center, Massachusetts General Hospital, Boston, MA; Division of Gastroenterology, University of Michigan, Ann Arbor, MI

## Abstract

**Background:** Statins may be protective in viral infection and have been proposed as treatment in severe acute respiratory syndrome coronavirus-2 (SARS-CoV-2) infection.

**Objective:** We evaluated the effect of statins on mortality in four groups hospitalized with (SARS-CoV-2) infection (continued statin, newly initiated statin, discontinued statin, never on statin).

**Design:** In a single center cohort study of 1179 patients hospitalized with SARS-CoV-2 infection, the outcome of death, Intensive Care Unit (ICU) admission or hospital discharge was evaluated. Patients’ statin use, laboratory data, and co-morbidities were determined via chart review and electronic health records. Using marginal structural models to account for timing of statin initiation and competing risks, we compared the likelihood of severe outcomes in the four statin exposure groups.

**Setting:** Academic medical center in the United States

**Participants:** Patients hospitalized with SARS-CoV-2 infection

**Measurements:** 28-day mortality, ICU admission, or discharge

**Results:** Among 1179 patients, 360 were never on a statin, 311 were newly initiated on a statin, 466 were continued on a statin, and 42 had a statin discontinued. In this cohort, 154 (13.1%) patients died by 28-days. With marginal structural model analysis, statin use reduced the hazard of 28-day mortality (HR 0.566 [CI 0.372, 0.862], p = 0.008). Both new initiation of statins (HR 0.493 [CI 0.253, 0.963], p=0.038) and continuing statin therapy reduced the hazard of 28-day mortality (HR 0.270 [CI 0.114, 0.637], p=0.003). Sensitivity analysis found that statin use was associated with improved mortality for patients > 65 years, but not for patients 65 years or younger.

**Limitation:** Observational design

**Conclusion:** Statin therapy during hospitalization for SARS-CoV-2 infection, including new initiation and continuation of therapy, was associated with reduced short-term mortality.

## Introduction

Severe acute respiratory syndrome coronavirus-2 (SARS-CoV-2) and the associated disease (COVID-19) has resulted in millions of deaths (1). During the initial rush for treatments, over 100 off-label and experimental drugs were used to treat patients with COVID-19 (2). Off-label statin use was considered early in the pandemic at Massachusetts General Hospital (MGH) for several reasons, including: reports of cardiac complications and death due to COVID-19 (3, 4) and the cardioprotective effect of statins (5, 6); statins are low cost, widely available and generally safe (7); and statins blunt the hyperinflammatory response from infection (8-10). It was also suggested that statins block SARS-CoV-2 infectivity via binding to the main protease mediating viral entry, inhibiting the virus’ ability to invade cells (11).

The safety and efficacy of statins for the treatment of patients hospitalized with COVID-19 has remained uncertain. Prior to the COVID-19 pandemic, the impact of statins in acute respiratory distress syndrome (ARDS) was unclear, with some trials showing no impact on mortality (12, 13), while others found a significant improvement (14, 15). Observational studies have similarly yielded mixed results regarding the effects of statins in COVID-19. These investigations have been limited by small sample size (16-19), insufficient adjustments for time varying confounders (16, 20, 21), and lack of a sub-cohort newly initiated on statins for COVID-19 (20, 22-24).

During the surge in Spring 2020, a multidisciplinary group of physicians at MGH created clinical guidelines that recommended starting statin therapy on patients hospitalized for COVID-19 (25, 26). Clinicians were advised to continue pre-hospital statins and to start atorvastatin 40 mg daily on patients who had an evidence-based indication (e.g. hyperlipidemia). At times, clinicians were also advised to start atorvastatin if the patient with COVID-19 had no contraindication, though this guidance changed over the study period. This placed MGH in a unique position to evaluate empirically the impact of starting patients on statin therapy during hospitalization due to COVID-19.

We aimed to evaluate the effect of statins on 28-day mortality in patients hospitalized with COVID-19, using robust causal inference methods (i.e., marginal structural Cox proportional hazards models) to account for variable timing of statin initiation and competing risks. Notably, our approach takes into account that the decision to initiate statins was influenced both by baseline confounding factors as well as patients’ changing health condition during hospitalization. We further aimed to evaluate the specific effect of new statin initiation on survival.

## Methods

### Patient Selection

This study is based on an MGH cohort described previously (27). Inclusion criteria included ≥ 18 years of age and confirmed SARS-CoV-2 infection with reverse transcriptase-polymerase chain reaction testing of nasopharyngeal or sputum specimens. For this study, we included patients hospitalized between March and June 2020. This study was approved by the local Institutional Review Board (IRB # 2020P000829); a waiver of informed consent was granted.

### Statin Exposure

Patients were categorized into four statin groups: (A) continued home statin during hospitalization (“continued”), (B) discontinued home statin during hospitalization (“discontinued”), (C) newly initiated statin during hospitalization (“newly initiated”), and (D) did not use statins during or prior to hospitalization (“never”) [**Figure 1]**. Manual chart review, supplemented by extracted medication orders from electronic health records in the Mass General Brigham Enterprise Data Warehouse (EDW), was performed to assess statin use both prior to and during hospitalization.

**Figure 1.**
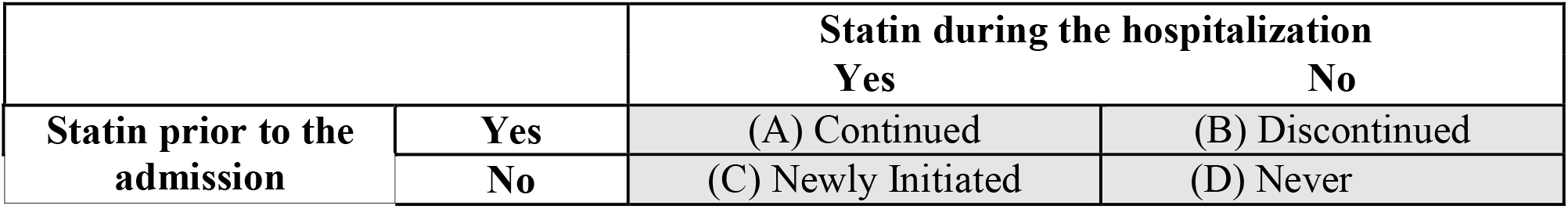
Definition of statin treatment groups.

### Covariates and Outcomes

Chart abstraction collected information on patient age, sex, smoking status, medical comorbidities, medications, hospitalization date, date of ICU admission, date of death or discharge, need for invasive mechanical ventilation, renal replacement therapy, and medications used at hospitalization. Laboratory values, self-reported race/ethnicity and body mass index (BMI) were extracted from the electronic health record.

Patients were followed from hospitalization until death, discharge, or 28 days from presentation to care. The primary outcome was time to death. Patients who were discharged to palliative care were classified as deceased on the day of their discharge. The secondary outcome was time to a composite outcome of ICU admission or death. For this secondary outcome, a patient must have received statins during hospitalization but prior to ICU admission to be categorized as newly initiated or continued.

### Statistical Analysis

We examined the differences between baseline characteristics among the four statin treatment groups. The median and interquartile ranges are presented for continuous variables and the counts and percentage are presented for categorical variables. For statistical tests, we conducted Fisher’s exact test for categorical variables and Kruskal-Wallis test by ranks for continuous variables.

Analyses that stratify patients by in-hospital statin use are subject to both immortal time bias and time-varying confounding, as a patient’s changing health condition affected when and whether they initiated statins (28). To address these challenges, we used marginal structural Cox proportional hazards models to evaluate the effect of statin initiation on the primary and secondary outcomes (29). Patients who were discharged alive or to a non-palliative care facility were categorized together as discharged. We fit a pooled multinomial regression to account for discharge as a competing risk alongside the outcome to fit the weighted Cox model (30). If patients did not experience the outcome and were not discharged by 28 days, they were treated as administratively censored.

Using the marginal structural Cox model, we estimated the hazard ratio of each outcome for initiating statins vs. not initiating statins by the previous day. We excluded patients who experienced the outcome on the day of admission, as there was no opportunity for in-hospital statin use to affect the risk of the outcome. The model we fit assumes that once initiated, the duration of in-hospital statin use does not affect the hazard of the outcome.

To fit the marginal structural Cox model, we used inverse probability weighting with stabilized weights trimmed at the 5^th^ and 95^th^ percentiles. Because all patients were followed until 28 days unless they experienced the outcome or were discharged, censoring weights were not used. The following baseline variables were included in the weighting models: demographic variables (sex, age>65 years, race, active smoker, BMI≥30), comorbidities on admission (coronary artery disease, congestive heart failure, hypertension, diabetes, dyslipidemia, chronic liver disease, active cancer, pulmonary disease), ACE inhibitor medication status at presentation to care, number of days from March 1^st^, 2020 to the date of hospitalization to account for era effect, and prior statin usage. The following time-varying daily lab measurements were also included and log-transformed: absolute lymphocyte count, white blood cell count (WBC), aspartate aminotransferase (AST), C-reactive protein (CRP), creatine kinase (CK) and alanine aminotransferase (ALT). To account for missingness in both baseline and time-varying data, we used multiple imputation with 25 imputations. Peak lab values were calculated for each statin group, defined as the highest lab measurement observed between hospital admission and death, discharge, or end of follow-up, whichever came first. These lab values were calculated irrespective of the exact timing of statin initiation.

For the primary outcome of mortality, we adjusted for ICU admission status each day as a time-varying covariate. 95% confidence intervals and p-values were calculated, accounting for the uncertainty due to estimation of the weighting models (30), and results across imputations were combined using Rubin’s rules (31).

The analysis was repeated for the primary and secondary outcomes on the subset of patients who were not prior users (“Newly initiated” vs. “Never”) and patients who were prior users (“Continued” vs. “Discontinued”). In total, six models were fit. An additional analysis was performed using E-values to assess robustness of the observed results to unmeasured confounding. The E-value is a measure that assesses how strong an unmeasured confounder would have to be to fully explain away the observed results (32, 33).Stratified analysis was also performed to assess whether the effect of statins in patients above and below 65 years of age. All analyses were conducted in R Version 4.0.2 (34). The *nnet* package (35) was used to fit the multinomial regression model and the *Jomo* package (36) was used to perform multiple imputation of multilevel data, which included demographic characteristics as well as time-varying laboratory measures.

## Results

### Baseline Characteristics

Overall, 1179 adult patients with confirmed SARS-CoV-2 infection were included, after the exclusion of 7 patients who were deceased, discharged, or censored on the day of their hospital admission **(Appendix Table 1)**. 676 (57.3%) were male, 443 (37.6%) were at least 65 years old, and 493 (45.9%) had a BMI ≥30. Patient characteristics differed by statin group **(Table 1)**. Patients on statins prior to hospitalization (groups A and B) were older (median age 69 vs. 52 years, p < 0.001), and had higher rates of coronary artery disease (28.7% vs 3.1%, p < 0.001), congestive heart failure (18.9% vs. 5.1%, p < 0.001), hypertension (74.4% vs. 34.3%, p < 0.001), type 2 diabetes (55.9% vs. 17.1%, p < 0.001), and dyslipidemia (66.7% vs. 15.6%, p < 0.001) than those not on statins prior to hospitalization (groups C and D). White/non-Hispanics were more likely to be on statins prior to hospitalization than Hispanics (56.4% of White/non-Hispanics on statins vs. 32.9% of Hispanics on statins, p < 0.001).

**Table 1:** Patient Characteristics at Hospitalization for COVID-19. Stratified by (A) continued statin during hospitalization (Continued), (B) discontinued statin during hospitalization (Discontinued), (C) newly initiated statin during hospitalization (Newly Initiated), and (D) no statin prior to admission nor during hospitalization (Never).

| Total 1,179 subjects | (A) Continued<br>(n=466) | (B) Discontinued<br>(n=42) | (C) Newly<br>Initiated<br>(n=311) | (D) Never<br>(n=360) | p-value* |
| --- | --- | --- | --- | --- | --- |
| **Demographics** |  |  |  |  |  |
| **Male sex, n/N (%)** | 298/466 (63.9%) | 26/42 (61.9%) | 173/311 (55.6%) | 179/360 (49.7%) | 0.001 |
| **Age, median (IQR)** | 68 (59-78) [n=466] | 70 (58-78) [n=42] | 55 (43-66) [n=311] | 48 (35-62) [n=360] | <0.001 |
| **Age > 65, n/N (%)** | 265/466 (56.9%) | 24/42 (57.1%) | 83/311 (26.7%) | 71/360 (19.7%) | <0.001 |
| **Race/Ethnicity, n/N (%)** |  |  |  |  |  |
| 1. White | 233/457 (51%) | 23/42 (54.8%) | 84/301 (27.9%) | 114/355 (32.1%) | <0.001 |
| 2. Black or African American | 40/457 (8.8%) | 7/42 (16.7%) | 43/301 (14.3%) | 37/355 (10.4%) |  |
| 3. Hispanic | 131/457 (28.7%) | 10/42 (23.8%) | 129/301 (42.9%) | 159/355 (44.8%) |  |
| 4. Other (American Indian, Alaska Native, Asian, Native Hawaiian, Pacific Islander, others) | 53/457 (11.6%) | 2/42 (4.8%) | 45/301 (15%) | 45/355 (12.7%) |  |
| **Active smoker, n/N (%)** | 24/442 (5.4%) | 7/37 (18.9%) | 15/275 (5.5%) | 39/330 (11.8%) | <0.001 |
| **BMI, median (IQR)** | 29 (26-34) [n=430] | 29 (25-32) [n=38] | 30 (26-34) [n=290] | 29 (25-34) [n=316] | 0.735 |
| **BMI >= 30, n/N (%)** | 192/430 (44.7%) | 14/38 (36.8%) | 141/290 (48.6%) | 146/316 (46.2%) | 0.500 |
| **Comorbidities on Admission, n/N (%)** |  |  |  |  |  |
| **Coronary Artery Disease** | 140/466 (30%) | 6/42 (14.3%) | 9/311 (2.9%) | 12/360 (3.3%) | <0.001 |
| **Congestive Heart Failure** | 90/466 (19.3%) | 6/42 (14.3%) | 15/311 (4.8%) | 19/360 (5.3%) | <0.001 |
| **Hypertension** | 349/466 (74.9%) | 29/42 (69%) | 130/311 (41.8%) | 100/360 (27.8%) | <0.001 |
| **Diabetes** | 261/466 (56%) | 24/42 (57.1%) | 76/311 (24.4%) | 42/360 (11.7%) | <0.001 |
| **Dyslipidemia** | 320/466 (68.7%) | 19/42 (45.2%) | 59/311 (19%) | 46/360 (12.8%) | <0.001 |
| **Chronic kidney disease** | 121/458 (26.4%) | 9/41 (22%) | 31/299 (10.4%) | 32/356 (9%) | <0.001 |
| Dialysis | 18/458 (3.9%) | 1/41 (2.4%) | 7/299 (2.3%) | 6/356 (1.7%) | 0.238 |
| **Chronic liver Disease\*\*** | 37/458 (8.1%) | 5/41 (12.2%) | 30/299 (10%) | 36/353 (10.2%) | 0.567 |
| Alcohol-related Cirrhosis | 4/458 (0.9%) | 2/41 (4.9%) | 5/299 (1.7%) | 4/353 (1.1%) | 0.161 |
| NAFLD | 22/458 (4.8%) | 0/41 (0%) | 16/299 (5.4%) | 14/353 (4%) | 0.499 |
| **Current Viral Hepatitis** | 18/466 (3.9%) | 1/42 (2.4%) | 11/311 (3.5%) | 25/360 (6.9%) | 0.125 |
| **HIV** | 6/413 (1.5%) | 0/40 (0%) | 4/283 (1.4%) | 6/329 (1.8%) | 0.947 |
| **History of Cancer** | 88/459 (19.2%) | 10/42 (23.8%) | 43/298 (14.4%) | 32/354 (9%) | <0.001 |
| **Pulmonary Disease** | 167/461 (36.2%) | 14/41 (34.1%) | 76/301 (25.2%) | 91/357 (25.5%) | 0.001 |
| COPD | 71/461 (15.4%) | 6/41 (14.6%) | 20/301 (6.6%) | 23/357 (6.4%) | <0.001 |
| Asthma | 60/461 (13%) | 7/41 (17.1%) | 37/301 (12.3%) | 55/357 (15.4%) | 0.548 |
| ILD | 3/461 (0.7%) | 0/41 (0%) | 3/301 (1%) | 3/357 (0.8%) | 0.938 |
| Home oxygen supplementation | 14/461 (3%) | 1/41 (2.4%) | 3/301 (1%) | 3/357 (0.8%) | 0.065 |
| **Obstructive Sleep Apnea (OSA)** | 46/466 (9.9%) | 4/42 (9.5%) | 16/311 (5.1%) | 12/360 (3.3%) | 0.001 |
| **History of organ transplant** | 13/462 (2.8%) | 2/41 (4.9%) | 6/301 (2%) | 7/356 (2%) | 0.480 |
| **Treatment with immunosuppressing agent in the past 6 months** | 36/448 (8%) | 4/38 (10.5%) | 16/295 (5.4%) | 22/353 (6.2%) | 0.366 |
| **Medications at Presentation to Care, n/N (%)** |  |  |  |  |  |
| **Type and dose of statin (at PTC)** |  |  |  |  |  |
| Atorvastatin | 309/458 (67.5%) | 28/41 (68.3%) |  |  | 0.358 |
| Atorvastatin 10mg-20mg | 95/309 (30.7%) | 7/28 (25%) |  |  |  |
| Atorvastatin 40mg-60mg | 116/309 (37.5%) | 12/28 (42.9%) |  |  |  |
| Atorvastatin 80mg | 85/309 (27.5%) | 7/28 (25%) |  |  |  |
| Unknown Dosage | 13/309 (4.2%) | 2/28 (7.1%) |  |  |  |
| Rosuvastatin | 39/458 (8.5%) | 1/41 (2.4%) |  |  |  |
| Other Statins | 110/458 (24%) | 12/41 (29.3%) |  |  |  |
| **Type and dose of statin (during hospitalization)** |  |  |  |  |  |
| Atorvastatin | 331/434 (76.3%) |  | 274/285 (96.1%) |  | <0.001 |
| Atorvastatin 10mg-20mg | 76/331 (23%) |  | 53/274 (19.3%) |  |  |
| Atorvastatin 40mg-60mg | 138/331 (41.7%) |  | 147/274 (53.6%) |  |  |
| Atorvastatin 80mg | 117/331 (35.3%) |  | 74/274 (27%) |  |  |
| Unknown Dosage | 0/331 (0%) |  | 0/274 (0%) |  |  |
| Rosuvastatin | 33/434 (7.6%) |  | 6/285 (2.1%) |  |  |
| Other Statins | 70/434 (16.1%) |  | 5/285 (1.8%) |  |  |
| **Duration of pre-admission statin use at PTC** |  |  |  |  |  |
| <= 1 year | 54/466 (11.6%) | 4/42 (9.5%) |  |  | 0.973 |
| >1 year | 244/466 (52.4%) | 23/42 (54.8%) |  |  |  |
| Unknown | 168/466 (36.1%) | 15/42 (35.7%) |  |  |  |
| **ACE Inhibitor** | 116/466 (24.9%) | 4/42 (9.5%) | 30/311 (9.6%) | 33/360 (9.2%) | <0.001 |
| **Azithromycin** | 17/457 (3.7%) | 2/42 (4.8%) | 18/302 (6%) | 22/353 (6.2%) | 0.326 |
| **Immunosuppressants** | 42/466 (9%) | 3/42 (7.1%) | 20/311 (6.4%) | 22/360 (6.1%) | 0.396 |
| Oral Steroid | 29/466 (6.2%) | 3/42 (7.1%) | 13/311 (4.2%) | 17/360 (4.7%) | 0.512 |
| **Immunomodulators** | 13/466 (2.8%) | 2/42 (4.8%) | 10/311 (3.2%) | 8/360 (2.2%) | 0.598 |
| Hydroxychloroquine | 4/466 (0.9%) | 0/42 (0%) | 3/311 (1%) | 3/360 (0.8%) | 1.000 |
| **Presentation to Care, n/N (%)** |  |  |  |  |  |
| **Symptomatic (yes/no)** | 439/462 (95%) | 36/40 (90%) | 305/309 (98.7%) | 333/357 (93.3%) | 0.001 |
| **First Available Labs, median (IQR)\*\*\*** |  |  |  |  |  |
| White blood cell count, /μL | 6.5 (4.9-8.3) [n=466] | 7.1 (5.2-10.5) [n=40] | 6.8 (5.2-9.2) [n=311] | 6.7 (4.9-9.2) [n=349] | 0.152 |
| Aspartate aminotransferase, U/L | 40 (28-56) [n=463] | 58 (30-121) [n=38] | 44 (31-65) [n=311] | 48 (30-72) [n=338] | <0.001 |
| Alanine aminotransferase, U/L | 27 (18-42) [n=463] | 34 (23-62) [n=38] | 34 (21-58) [n=311] | 34 (21-66) [n=338] | <0.001 |
| Total bilirubin, mg/dL | 0.5 (0.4-0.7) [n=463] | 0.5 (0.4-0.8) [n=38] | 0.5 (0.4-0.7) [n=311] | 0.5 (0.3-0.6) [n=339] | 0.125 |
| Alkaline phosphatase, U/L | 86 (67-108) [n=463] | 91 (69-127) [n=38] | 75 (59-95) [n=311] | 80 (62-108) [n=338] | <0.001 |
| Troponin, ng/L | 17 (8-39) [n=451] | 39 (14-90) [n=38] | 8 (6-17) [n=305] | 7 (6-18) [n=322] | <0.001 |
| C-reactive protein, mg/L | 74 (33-142) [n=459] | 85 (48-152) [n=36] | 89 (49-152) [n=311] | 68 (31-136) [n=330] | 0.001 |
| Erythrocyte sedimentation rate, mm/h | 41 (27-63) [n=426] | 49 (35-84) [n=32] | 42 (28-65) [n=297] | 35 (21-54) [n=295] | <0.001 |
| Creatine kinase, U/L | 114 (63-217) [n=461] | 298 (82-702) [n=36] | 114 (67-223) [n=310] | 132 (65-349) [n=331] | 0.003 |
| D-dimer, ng/mL | 1063 (695-1812) [n=455] | 1554 (826-6580) [n=37] | 1074 (693-1770) [n=305] | 949 (608-1661) [n=329] | 0.002 |
| Absolute lymphocyte count, K/μL | 0.9 (0.6-1.3) [n=466] | 1 (0.6-1.5) [n=40] | 1 (0.7-1.3) [n=311] | 1 (0.7-1.4) [n=341] | 0.132 |
| **Calendar Days Since 03/01/2020, median (IQR)** |  |  |  |  |  |
| Number of days from 03/01/2020 to hospital admission date | 43 (33-52) [n=466] | 50 (41-56) [n=42] | 37 (30-44) [n=311] | 46 (40-54) [n=360] | <0.001 |

Non-alcoholic Fatty Liver Disease (NAFLD), HIV (Human Immunodeficiency Virus), ILD (interstitial lung disease), COPD (chronic obstructive pulmonary disease)
\* p-values were calculated comparing all 4 groups using the Kruskal-Wallis test for continuous variables and Fisher's exact test for categorical variables.
\*\* Unknown and other chronic liver disease not presented in the table
\*\*\* Note that this is first available lab result following admission

In total, 777 patients (65.9%) received a statin during their hospitalization for COVID-19. Atorvastatin was the most common statin prescribed before and during hospitalization. 274/285 (96.1%) patients newly initiated on statins and 331/434 (76.3%) patients continued on statins were prescribed atorvastatin. The most common dosage was moderate intensity (atorvastatin 40mg daily). Among those continued on statins, the majority were prescribed statins for more than a year prior to hospitalization (244/466, 52.4%). **Appendix Table 2** contains information on when statins were initiated during hospitalization relative to admission date.

According to first laboratory measurements obtained during hospitalization, the group of patients who discontinued statins at hospitalization had higher erythrocyte sedimentation rate (p = 0.046), CK (p = 0.001), troponin (p < 0.001), and D-dimer (p = 0.002) compared to the other cohorts **(Table 1)**.

### Unadjusted Analysis

In the complete cohort, 154 (13.1%) patients died and 841 (71.3%) were discharged within 28 days of follow up. In unadjusted analyses, patients on statins during hospitalization (groups A and C) had similar rates of death (108 (13.9%) vs. 46 (11.4%), p = 0.273), but higher rates of ongoing hospitalization at 28 days (144 (18.5%) vs. 40 (10.0%), p < 0.001) and ICU admission (276 (35.5%) vs. 85 (21.1%), p < 0.001) than those not on statins during hospitalization (groups B and D) (**Table 2**).

**Table 2:** Unadjusted Patient Outcomes at 28 Days.

| Total 1,179 subjects | (A) Continued<br>(n=466) | (B)<br>Discontinued<br>(n=42) | (C) Newly<br>Initiated<br>(n=311) | (D) Never<br>(n=360) | P-value*<br>* |
| --- | --- | --- | --- | --- | --- |
| **28-day Outcomes from Presentation to Care** |  |  |  |  |  |
| **Patient status at 28 days, n (%)** |  |  |  |  |  |
| Deceased* | 78/466 (16.7%) | 14/42 (33.3%) | 30/311 (9.6%) | 32/360 (8.9%) | <0.001 |
| Discharged alive | 252/466 (54.1%) | 20/42 (47.6%) | 176/311 (56.6%) | 257/360 (71.4%) |  |
| Transfer to other facility (non-palliative care) | 61/466 (13.1%) | 4/42 (9.5%) | 36/311 (11.6%) | 35/360 (9.7%) |  |
| Still hospitalized at 28 days after presentation to care | 75/466 (16.1%) | 4/42 (9.5%) | 69/311 (22.2%) | 36/360 (10.0%) |  |
| Days from hospital admission to death, median (IQR) | 10 (6-15) [n=78] | 3 (1-4) [n=14] | 12 (7-16) [n=30] | 6 (3-13) [n=32] | <0.001 |
| Days from hospital admission to discharge or transfer to non-palliative facility, median (IQR) | 7 (4-10) [n=313] | 4 (1-6) [n=24] | 7 (4-10) [n=212] | 5 (3-8) [n=292] | <0.001 |
| **Patient events during 28 days of follow-up, n (%)** |  |  |  |  |  |
| ICU admission | 145/466 (31.1%) | 8/42 (19.0%) | 131/311 (42.1%) | 77/360 (21.4%) | <0.001 |
| Invasive intubation | 122/466 (26.2%) | 6/42 (14.3%) | 110/311 (35.4%) | 65/360 (18.1%) | <0.001 |
| Bacterial Pneumonia | 132/466 (28.3%) | 9/42 (21.4%) | 106/311 (34.1%) | 71/360 (19.7%) | <0.001 |
| Acute Respiratory Distress Syndrome | 112/466 (24.0%) | 5/42 (11.9%) | 105/311 (33.8%) | 61/360 (16.9%) | <0.001 |
| Stroke / Cerebrovascular accident | 3/466 (0.6%) | 1/42 (2.4%) | 3/311 (1.0%) | 2/360 (0.6%) | 0.390 |
| Cardiac arrest | 2/466 (0.4%) | 1/42 (2.4%) | 2/311 (0.6%) | 2/360 (0.6%) | 0.390 |
| Rhabdomyolysis/myositis | 18/466 (3.9%) | 2/42 (4.8%) | 18/311 (5.8%) | 21/360 (5.8%) | 0.499 |
| Liver dysfunction | 80/466 (17.2%) | 11/42 (26.2%) | 71/311 (22.8%) | 64/360 (17.8%) | 0.123 |

IQR (Interquartile range)
\*Deceased includes 3 patients who were discharged to hospice care
\*\* p-values were calculated comparing all 4 groups using the Kruskal-Wallis test for continuous variables and Fisher's exact test for categorical variables.

### Peak Laboratory Values

Unadjusted peak liver biochemistries and inflammatory markers differed across statin groups (**Table 3**), with the highest peaks of AST and ALT in the newly initiated statin group. Patients newly initiated on statins had a lower peak CK than those who discontinued statins (median and interquartile range: 222 [90-636] U/L vs. 374 [89-838] U/L), but higher peak CK than those who continued (178 [85-507] U/L) or were never on statins (173 [78-578] U/L).

**Table 3:** Unadjusted Peak Laboratory Values by Statin Group.

| Total 1,179 subjects | (A) Continued<br>(n=466) | (B)<br>Discontinued<br>(n=42) | (C) Newly<br>Initiated<br>(n=311) | (D) Never<br>(n=360) | p-value* |
| --- | --- | --- | --- | --- | --- |
| **Peak Labs, median**<br>**(IQR)** |  |  |  |  |  |
| White blood cell<br>count, /μL | 8.7 (6.6-12.5)<br>[n=466] | 7.9 (6.3-16)<br>[n=40] | 9.9 (7.1-15.2)<br>[n=311] | 8.3 (6.2-12.1)<br>[n=349] | 0.001 |
| Aspartate<br>aminotransferase, U/L | 65 (40-119)<br>[n=463] | 60 (32-208)<br>[n=38] | 84 (45-150)<br>[n=311] | 66 (35-127)<br>[n=338] | 0.008 |
| Alanine<br>aminotransferase, U/L | 46 (26-85)<br>[n=463] | 47 (23-121)<br>[n=38] | 68 (31-120)<br>[n=311] | 57 (26-118)<br>[n=338] | <0.001 |
| Total bilirubin, mg/dL | 0.7 (0.5-1.1)<br>[n=463] | 0.5 (0.4-0.8)<br>[n=38] | 0.6 (0.5-1)<br>[n=311] | 0.6 (0.4-0.9)<br>[n=339] | 0.011 |
| Alkaline phosphatase,<br>U/L | 105 (79-166)<br>[n=463] | 101 (77-155)<br>[n=38] | 97 (72-156)<br>[n=311] | 96 (72-140)<br>[n=338] | 0.017 |
| Troponin, ng/L | 23 (10-52)<br>[n=451] | 40 (14-90)<br>[n=38] | 12 (6-29)<br>[n=305] | 8 (6-23)<br>[n=322] | <0.001 |
| C-reactive protein,<br>mg/L | 145 (69-252)<br>[n=459] | 141 (50-231)<br>[n=36] | 151 (82-283)<br>[n=311] | 117 (52-175)<br>[n=330] | <0.001 |
| Erythrocyte<br>sedimentation rate,<br>mm/h | 57 (36-104)<br>[n=426] | 68 (36-90)<br>[n=32] | 72 (41-114)<br>[n=297] | 44 (26-74)<br>[n=295] | <0.001 |
| Creatine kinase, U/L | 178 (85-507)<br>[n=461] | 374 (89-838)<br>[n=36] | 222 (90-636)<br>[n=310] | 173 (78-578)<br>[n=331] | 0.028 |
| D-dimer, ng/mL | 1863 (986-3596) [n=455] | 1944 (1075-7380) [n=37] | 2090 (1038-5098) [n=305] | 1241 (713-3270) [n=329] | <0.001 |
| Absolute lymphocyte<br>count, K/μL | 1.6 (1.1-2.2)<br>[n=466] | 1.5 (1-2.5)<br>[n=40] | 1.9 (1.4-2.4)<br>[n=311] | 1.7 (1.3-2.3)<br>[n=341] | <0.001 |

\* p-values were calculated comparing all 4 groups using the Kruskal-Wallis test for continuous variables and Fisher's exact test for categorical variables.

### Primary Outcome Analysis

The median time to death in each statin group can be found in **Table 1**. Overall, statin usage during hospitalization (groups A and C) decreased the hazard of death (HR 0.566 [95% CI 0.372, 0.862], p=0.008). In the sub-group of patients *not on statin* therapy prior to hospitalization (groups C and D), new statin initiation at hospitalization (group C) decreased the hazard of death (HR 0.493 [95% CI 0.253, 0.963], p=0.038). The sub-group of patients *on statin* therapy prior to hospitalization (groups A and B), continued statin usage (group A) also decreased the hazard of death (HR 0.270 [95% CI 0.114,0.637], p=0.003) (**Figure 2)**. A summary of the distribution of missing laboratory measurements can be found in **Appendix Table 3**.

**Figure 2:**
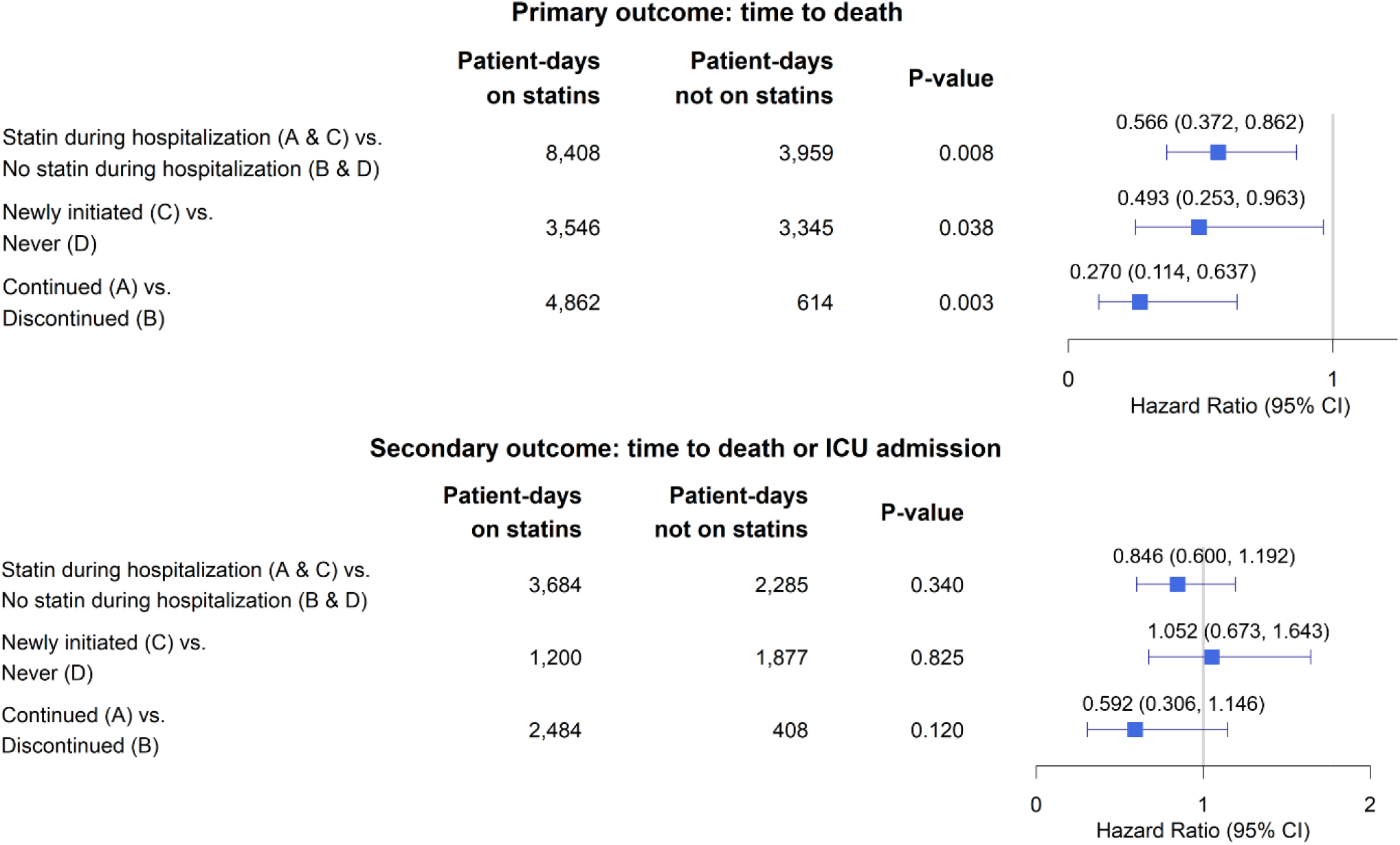
Marginal Structural Model Outputs for Primary and Secondary Outcomes. Estimates were obtained from fitting marginal structural Cox models adjusted for the following baseline covariates: sex, age>65 years, race, active smoker, BMI≥30, comorbidities on admission (coronary artery disease, congestive heart failure, hypertension, diabetes, dyslipidemia, chronic liver disease, active cancer, pulmonary disease), ACE inhibitor use, number of days since March 1st, 2020, and prior statin usage. The following time-varying covariates were adjusted for as well: ALC, WBC, AST, CRP, CK, ALT, and ICU admission status. Models were fit accounting for immortal time bias, time-varying confounding, and discharge as a competing risk.

### Secondary Outcome Analysis

To fit the marginal structural model for the secondary outcome of time to death or ICU admission, 198 patients were excluded because they were admitted to the ICU on their day of hospitalization and therefore did not contribute time at risk for the secondary outcome (**Appendix Table 1**). Of the 981 patients remaining, 588 (59.9%) had recorded statin use during their hospitalization. Statin use during hospitalization (groups A and C) did not change the hazard of the composite outcome of death or ICU admission (HR 0.846 [95% CI 0.600,1.192], p=0.340) (**Figure 2** and **Appendix Table 4**).

### Analysis of Unmeasured Confounding

For the primary outcome assessing mortality, the point estimate and the upper confidence limit of the E-value associated with statin use during hospitalization is 2.326 and 1.455, respectively. Given that the E-value of 2.326 is much greater than any observed known risk factors examined in the current study (with the exception of age), it is unlikely that an unmeasured confounder exists that would explain away the observed effect in the present analysis. The point estimate and the upper confidence limit of the E-value associated with new initiation of statins is 2.639 and 1.191, respectively. The point estimate and the upper confidence limit of the E-value associated with continuation of statins is 4.305 and 2.073, respectively.

### Sensitivity Analyses for Age

We fit a marginal structural model for the primary outcome of time to death in both a subgroup of patients 65 years of age or younger (746 patients) and patients older than 65 years (443 patients). Statin use during hospitalization did not change the hazard of death in patients ≤ 65 years (HR 1.175 [95% CI 0.520, 2.655], p=0.699) (**Appendix Table 5**); however, patients > 65 years were found to have a decreased hazard of death with any statin use during hospitalization (HR 0.477 [95% CI 0.292, 0.78], p=0.003) and within the subgroup of patients newly initiated on statins during hospitalization (HR 0.321 [ 95% CI 0.137, 0.752], p= 0.009) **(Appendix Table 6)**.

## Discussion

In this large cohort from a single tertiary medical center, we found that statin use during hospitalization for COVID-19 was associated with improved short-term mortality. The survival benefit was seen in those who continued statin therapy, as well as those who newly initiated therapy while hospitalized. A sensitivity analysis found that statin use was associated with improved mortality for patients older than 65 years, but not for patients 65 years old or younger.

Our study confirms and expands on prior work. A recent propensity score-matched analysis found that statin use prior to hospitalization reduces the risk of short-term in-hospital mortality from COVID-19 (23). Our study probed further, into whether in-hospital statin use had a similar effect on mortality. To investigate the impact of statins administered during hospitalization, we used marginal structural models, which account for both survivorship bias (i.e., patients need to survive long enough to begin statins) and time-varying confounding bias (i.e., patient health status during hospitalization changes over time, affecting the propensity of initiating treatment). Our study accounted for a wide variety of time-varying confounders, which we believe accurately captures shifts in the propensity of initiating treatment. We further expanded on prior work by specifically evaluating the effect of statin initiation during hospitalization without prior use, because 26% of our cohort was newly initiated on statins during COVID-19 hospitalization. Overall, the novel findings presented here are that statin therapy during hospitalization, whether it be a new or continued prescription, was associated with improved mortality.

The composition of our cohort was similar to other published patient databases of patients with COVID-19 (17, 22, 24). The median age of patients was 60 years old (IQR 47-73 years). Obese patients with a BMI ≥30 represented almost half the population, consistent with evidence that obesity is a risk factor for hospitalization with COVID-19 (37). Racial and ethnic demographics vary immensely across the literature. In this study, White/Non-Hispanic (38.5%) and Hispanic patients (36.4%) were most common, followed by Black/Non-Hispanic patients (10.8%). This cohort’s mortality rate (13.1%) was similar to the mean published United States hospital mortality rates for patients admitted with COVID-19 during the first 6 months of the pandemic (11.8%) among 955 hospitals (38).

Our findings contradict some prior reports. One meta-analysis of 3,449 patients in 9 observational studies found that statin use did not improve severe outcomes or mortality in COVID-19 (39). The majority of these studies were small in size (as few as 50 patients) and most failed to control for potential confounders. Comorbidities such as diabetes, cardiovascular disease and obesity are established risk factors for more severe COVID-19 disease, and patients prescribed statins are more likely to have these comorbidities, highlighting the critical need to control for confounding in this analysis (18). Additionally, most prior work either examined the relationship with antecedent statin usage prior to hospitalization (19, 21-24) or exclusively during hospitalization (16, 18, 20). Our study had several advantages over prior reports: our study investigated the influence of statin use both prior to and during hospitalization, minimized immortal time bias, adjusted for time-varying confounders, and minimized era effect (or the potential changes in COVID-19 care over time). These advantages provide clarity not offered by prior work.

The duration of statin therapy required to provide mortality benefit in COVID-19 remains unclear. This study suggests that statins are associated with a survival benefit even after brief exposure during hospitalization. Interestingly, in a large trial measuring mortality in US Veterans over the age of 75, statins did not affect mortality from cardiovascular events until at least 2 years of daily usage (40). However, a cardiology trial found that administration of atorvastatin within 24 to 96 hours of hospitalization for myocardial ischemia, compared with placebo, reduced the incidence of cardiovascular events including death, further myocardial infarction, and cardiac arrest (41). The main therapeutic mechanism of action of statins may be endothelial stabilization as well as its effects on inflammation (41). Given the association between COVID-19 infection and a hyperinflammatory response that provokes cardiovascular disorders (6), it is possible that new initiation of statins even for a short duration could have provided a positive impact on both mortality and secondary complications.

Another proposed explanation for statins’ mechanism of action in COVID-19 is the drug’s ability to inhibit hydroxymethylglutaryl-coenzyme A reductase, which may interfere with the virus’ invasion into cells by compromising the lipid-rich membrane required for SARS-CoV-2 to interact with the cellular receptor angiotensin-converting enzyme 2 (42, 43). If statins improve COVID-19 outcomes by inhibiting viral cell invasion, the benefit may be enhanced when statins are prescribed before infection.

In this study, we found an association with mortality benefit with statin use in adults over 65 years old but not in patients 65 years or younger. Given these findings were generated from a sensitivity analysis, we hesitate to provide age cut-offs for statin initiation for the treatment of COVID-19, but do not believe that age should be a contraindication to statin use during COVID-19 hospitalization.

Liver biochemistries are frequently elevated in severe COVID-19 disease and are associated with worse clinical outcomes (44). Separately, statins are known to increase liver biochemistries in certain patients (42). In our study, newly initiated statin use was associated with higher levels of peak AST and ALT throughout hospitalization; however, only 16% of patients on statins developed an ALT level greater than 5 times the upper limit of normal, which is a similar rate compared other cohorts with severe COVID (45). These findings are limited in that peak levels could have occurred before or after statin initiation and were unadjusted for confounding. Despite these limitations, there is no clear evidence that statin exposure during infection is associated with clinically important hepatotoxicity.

Myotoxicity is a known, albeit rare, complication of statins. In the context of COVID-19, where creatinine kinase elevations are prevalent, there is concern that statins could increase myotoxicity and subsequent creatinine kinase-induced nephrotoxicity. This was not seen among individuals newly initiated on statins or continued on statin therapy, with creatinine kinase peak levels never reaching three times the upper limit of normal, clinically considered the threshold for acute kidney injury due to pigment associated nephropathy (46).

Our findings must be interpreted in the context of study design. Although we utilized two data sources to confirm demographics, medications, laboratory data and clinical outcomes, misclassification errors are possible. We limited this error rate by employing physician review of discrepant data. Additionally, we were unable to account for patients who had dose escalations in their statin usage or who had their statin temporarily held during hospitalization. It is important to note that peak liver biochemistry levels (AST and ALT) as well as peak CK level were compared with unadjusted analyses and timing of statin initiation was not taken into account. Finally, patients who were on statin therapy prior to hospitalization but had the medication discontinued during hospitalization, presumably due to organ dysfunction, are a unique sub-cohort of patients that warrant further exploration. We utilized matched weighting within our marginal structural model to adjust for potential bias introduced by the extreme illness of this group.

After adjusting for cofounding factors, our study found that inpatient statin therapy was associated with improved 28-day mortality in COVID-19. This benefit was found in those who continued and newly initiated statins. Given the safety and availability of statins worldwide, a randomized controlled trial of statins in COVID-19 should be considered.

## Data Availability

This data was obtained from the MGH Covid-19 Data registry. Readers can contact the corresponding author for further information.

## Abbreviations

SARS-CoV-2: Severe Acute Respiratory Syndrome Coronavirus 2
COVID-19: Coronavirus disease 2019
MGH: Massachusetts General Hospital
ACE-2: Angiotensin-converting enzyme-2
AST: Aspartate Aminotransferase test
ALT: Alanine Aminotransferase test
ICU: Intensive Care Unit
HR: Hazard Ratio
CI: Confidence Interval
ARDS: acute respiratory distress syndrome
EDW: Enterprise Data Warehouse
BMI: Body Mass Index
WBC: white blood cell count
CRP: C-reactive protein
HIV: Human Immunodeficiency Virus
COPD: Chronic Obstructive Pulmonary Disease
ILD: Interstitial Lung Disease
IQR: Interquartile range
NAFLD: Non-alcoholic Fatty Liver Disease
ALC: Absolute lymphocyte count
CK: Creatine Kinase

