## Supplementary Tables for "Statins Are Associated with Improved 28-day Mortality in Patients Hospitalized with SARS-CoV-2 Infection"

**Supplementary Table 1**: Patients excluded from analyses due to experiencing the outcome, discharge, or censoring on the day of admission.

| **Exclusion event** | **Prior statin users** | **Non-prior statin users** |
| --- | --- | --- |
| Deceased | 4 | 0 |
| Discharged | 1 | 1 |
| Censored | 1 | 0 |
| Admitted to ICU | 78 | 120 |

*Note: Patients who were deceased, discharged, or censored on the day of admission were excluded from analyses of both primary and secondary outcomes. Patients were censored on the day of admission if admission occurred 28 days after presentation to care. Patients who were admitted to the ICU on the day of admission were excluded from analyses of the secondary outcome only.*

**Supplementary Table 2:** Patient counts, by number of days between admission and in-hospital statin initiation for patients who initiated statins prior to experiencing each outcome.

|  | **Patient counts** | |
| --- | --- | --- |
| **Days** | **Primary outcome** | **Secondary outcome** |
| 0 | 323 | 255 |
| 1 | 385 | 296 |
| 2 | 33 | 21 |
| 3 | 8 | 4 |
| 4 | 10 | 5 |
| 5 | 2 | 0 |
| 6 | 5 | 1 |
| 7 | 1 | 0 |
| 8 | 2 | 0 |
| 10 | 2 | 1 |
| 12 | 2 | 1 |
| 13 | 2 | 2 |
| 17 | 1 | 1 |
| 20 | 1 | 1 |

**Supplementary Table 3:** Completeness of laboratory measures used for estimation of inverse probability weights in the marginal structural model assessing the primary outcome, fit among all patients (mortality; A and C vs. B and D).

| **Laboratory test name** | **Patient-days with measured lab values (%)** | **Patient-days with prior lab values carried forward (%)** | **Patient-days with missing lab values that were multiply imputed (%)** |
| --- | --- | --- | --- |
| ALC | 79.6 | 13.0 | 7.4 |
| WBC | 84.9 | 8.5 | 6.6 |
| AST | 73.5 | 16.7 | 9.8 |
| CRP | 59.8 | 29.2 | 11 |
| CK | 61.0 | 27.3 | 11.7 |
| ALT | 73.5 | 16.7 | 9.8 |

*ALC: absolute lymphocyte count, WBC: white blood cell count, AST: aspartate aminotransferase, CRP: C-reactive protein, CK: creatine kinase, ALT: alanine aminotransferase.*

**Supplementary Table 4**: Secondary outcome rates and median time to secondary outcome, by statin usage group

| **Total:**  **981 subjects** | **(A) Continued**  **(n=374)** | **(B) Discontinued**  **(n=56)** | **(C)**  **Newly Initiated (n=214)** | **(D)**  **Never (n=337)** | **p-value** |
| --- | --- | --- | --- | --- | --- |
| Deceased or admitted to ICU, n (%) | 85 (22.7%) | 33 (58.9%) | 45 (21.0%) | 72 (21.4%) | <0.001 |
| Median days to death or ICU admission  (IQR) | 5 (2-9) | 1 (1-2) | 4 (2-7) | 2 (1-4) | <0.001 |

**Supplementary Table 5**: Marginal Structural Model Outputs for Primary and Secondary Outcomes, restricting to patients <= 65 years of age (736 patients). The same covariate adjustment approach was used as before.

|  |  | **Statin during hospitalization**  **(A & C)**  **vs.**  **No statin during hospitalization (B & D)** | **Newly initiated (C) vs.**  **Never (D)** | **Continued (A)**  **vs.**  **Discontinued (B)** |
| --- | --- | --- | --- | --- |
| Primary outcome: time to death | Hazard ratio (95% CI) | 1.175  (0.520, 2.655) | 1.619  (0.512, 5.124) | Model did not converge |
|  | P-value | 0.699 | 0.412 |  |
|  | Patient-days  on statins | 4,537 | 2,515 | 2,022 |
|  | Patient-days not on statins | 2,882 | 2,566 | 316 |
|  | Patient outcome rate, on statins | 23/429 (5.4%) | 10/228 (4.4%) | 13/201 (6.5%) |
|  | Patient outcome rate, not on statins | 9/307 (2.9%) | 7/289 (2.4%) | 2/18 (11.1%) |
| Secondary outcome: time to death or ICU admission | Hazard ratio (95% CI) | 1.043  (0.654, 1.665) | 1.346  (0.786, 2.305) | 0.804  (0.228, 2.831) |
|  | P-value | 0.859 | 0.278 | 0.734 |
|  | Patient-days  on statins | 1,619 | 754 | 865 |
|  | Patient-days not on statins | 1,586 | 1,383 | 203 |
|  | Patient outcome rate, on statins | 54/317 (17.0%) | 27/158 (17.1%) | 27/159 (17.0%) |
|  | Patient outcome rate, not on statins | 60/307 (19.5%) | 44/270 (16.3%) | 16/29 (55.2%) |

**Supplementary Table 6:**

Marginal Structural Model Outputs for Primary and Secondary Outcomes, restricting to patients > 65 years of age (443 patients). The same covariate adjustment approach was used as before.

|  |  | **Statin during hospitalization**  **(A & C)**  **vs.**  **No statin during hospitalization (B & D)** | **Newly initiated (C) vs.**  **Never (D)** | **Continued (A)**  **vs.**  **Discontinued (B)** |
| --- | --- | --- | --- | --- |
| Primary outcome: time to death | Hazard ratio (95% CI) | 0.477 (0.292, 0.78) | 0.321 (0.137, 0.752) | 0.224 (0.082, 0.617) |
|  | P-value | 0.003 | 0.009 | 0.004 |
|  | Patient-days  on statins | 3,871 | 1,031 | 2,840 |
|  | Patient-days not on statins | 1,077 | 779 | 298 |
|  | Patient outcome rate, on statins | 85/348 (24.4%) | 20/83 (24.1%) | 65/265 (24.5%) |
|  | Patient outcome rate, not on statins | 37/95 (38.9%) | 25/71 (35.2%) | 12/24 (50.0%) |
| Secondary outcome: time to death or ICU admission | Hazard ratio (95% CI) | 0.685 (0.408, 1.15) | 0.711 (0.253, 1.995) | 0.466 (0.177, 1.225) |
|  | P-value | 0.152 | 0.516 | 0.122 |
|  | Patient-days  on statins | 2,065 | 446 | 1,619 |
|  | Patient-days not on statins | 699 | 494 | 205 |
|  | Patient outcome rate, on statins | 76/271 (28.0%) | 18/56 (32.1%) | 58/215 (27.0%) |
|  | Patient outcome rate, not on statins | 45/94 (47.9%) | 28/67 (41.8%) | 17/27 (63.0%) |
